# The impact of central and obstructive respiratory events on cerebral oxygenation in adults with sleep disordered breathing

**DOI:** 10.1101/2023.01.05.23284218

**Authors:** Ramin Khatami, Dominik Gnaiger, Gordana Hügli, Ming Qi, Zhongxing Zhang

**Author notes:** Correspondence to: Zhongxing Zhang, PhD, Center for Sleep Medicine, Sleep Research and Epileptology, Clinic Barmelweid AG, 5017 Barmelweid, Switzerland.

## Abstract

Obstructive (OSA) and central sleep apnea (CSA) are two main types of sleep disordered breathing (SDB). While the changes in cerebral hemodynamics triggered by OSA events have been well studied using near-infrared spectroscopy (NIRS), they are essentially unknown in CSA in adults. Therefore, in this study we compared the changes in cerebral oxygenation between OSA and CSA events in adult patients using NIRS. Cerebral tissue oxygen saturation (StO2) in 13 severe SDB patients who had both CSA and OSA events was measured using frequency-domain NIRS. The changes in cerebral StO2 desaturation and blood volume (BV) in the first hour of natural sleep were compared between different types of respiratory events (i.e., 277 sleep hypopneas, 161 OSAs and 113 CSAs) with linear mixed-effect models controlling for confounders. All respiratory events occurred during non-rapid eye movement (NREM) sleep. We found that apneas events induced greater cerebral desaturations and BV fluctuations compared to hypopneas, but there was no difference between OSA and CSA. These results suggest that cerebral autoregulation in our patients are still capable to counteract the pathomechanisms of apneas, in particularly the negative intrathoracic pressure (ITP) caused by OSA events. Otherwise larger BV fluctuations in OSA compared to CSA should be observed due to the negative ITP that reduces cardiac stroke volume and leads to lower systematic blood supply. Our study suggests that OSA and CSA may have similar impact on cerebral oxygenation during NREM sleep in adult patients with SDB.

## 1. Introduction

Obstructive sleep apnea (OSA) and central sleep apnea (CSA) are two main types of sleep disordered breathing (SDB). OSA is the most common form of SDB with a prevalence of up to even about 40% in the general population^1^. OSA event is characterized by reduction or even cessation of breathing caused by an obstruction of the upper airway during sleep, usually at the level of the tongue and epiglottis due to a reduction in muscle tone^1, 2^. It can cause hypoxia (i.e., a decrease in blood oxygen saturation) and strong negative intrathoracic pressure (ITP) swings because the patients still have inhalation efforts in spite of the obstruction of upper airway. OSA event is usually terminated by arousal which is characterized by several seconds of wakefulness to regain muscle tone associated with a rise in heart rate and blood pressure. CSA is less common than OSA. CSA event is characterized by cessation of breathing efforts during sleep due to absent ventilatory drive (i.e., brain doesn’t send proper signals to the respiratory muscles) without upper airway obstruction^3, 4^. So unlike OSA, CSA usually cannot cause ITP swings as patients have no inhalation efforts. But similarly, the repetitive OSA and CSA events can both result in intermittent hypoxia, sleep fragmentations and recurrent arousals during sleep^5^. OSA is frequently associated with excessive daytime sleepiness (ESS) and neurocognitive dysfunction, and both OSA and CSA are high risk factors leading to the development of cardiovascular diseases including systemic hypertension, stroke and ischemic heart disease^6-10^. It is common to find OSA and CSA events in the same patient with SDB and patients who already have cardiovascular diseases such as heart failure^11^ and stroke^12^.

A better understanding of the changes in cerebral hemodynamics caused by OSA and CSA events may be the key to understand the acute (e.g., daytime sleepiness and bad mood) and chronic (e.g., hypertension and stroke) cerebral damage caused by SDB^10, 13-17^. In the past decades many studies have applied near-infrared spectroscopy (NIRS) to study the cerebral hemodynamic changes induced by OSA events^13, 18-22^ and even during treatment^17, 23, 24^ in patients. Compared to other neuroimaging methodologies such as functional magnetic resonance imaging(fMRI), positron emission tomography(PET), and single-photon emission computed tomography(SPECT), NIRS is naturally suitable to measure cerebral hemodynamics in human during all-night sleep because its sensors can be easily attached to human’s head and have good tolerance to movement^25, 26^. NIRS is sensitive to hemodynamic changes in microvascular bed (i.e., arterioles, venules and capillaries)^27, 28^. It can simultaneously measure changes in oxygenated hemoglobin (HbO2), de-oxygenated hemoglobin (HHb), blood volume (BV) and tissue oxygen saturation (StO2), thus providing better insights into the cerebral hemodynamic consequences induced by respiratory events.

Previous studies using NIRS have repeatedly shown a pattern of a decrease in HbO2 and StO2 while an increase in HHb during OSA events in patients^13, 17-24^. After the termination of the apneic event these parameters change in the opposite directions. However, very few NIRS data on CSA events have been reported so far. Several studies on cerebral hemodynamics in CSA events have been performed in infants especially in preterm infants^29-32^, because immaturity of their respiratory system is frequently manifested as CSA. The excessive or persistent apnea of prematurity during infancy is associated with significant falls in cerebral oxygenation^30, 32^, which is supposed to be associated with poor long term neurodevelopmental outcomes. However, infants have a general immaturity in many other systems such as cardiovascular and neuronal systems, making it complicated to interpret the cerebral hemodynamic changes in sleep apneas. For example, Jenni et al^31^ reported four different patterns of behavior of NIRS parameters during sleep apneas in preterm infants. Several recent studies also compared the cerebral StO2 desaturation between different types of sleep apneas in children with SDB but with inconsistent results. Tabone et al. found similar cerebral StO2 desaturations in CSA and OSA events and the decreases are significantly larger than the StO2 desaturations for sleep hypopneas^33^. However, Tamanyan et al. reported a greater cerebral StO2 desaturation in CSA events compared to obstructive events including the obstructive hypopneas and apneas^34^. It is remarkable that both studies reported that younger children have a greater cerebral desaturation compared to older ones. These results can be explained by a lower capacity of cerebrovascular autoreactivity and an immaturity of neural autonomic control in younger children^33, 34^. Therefore, the degree of NIRS cerebral desaturation in SDB may be age dependent and explain why results of infants and children cannot be applied to adults who have mature cerebrovascular and neural autonomic systems.

To the best of our knowledge, the differences in cerebral desaturations between CSA and OSA events in adults are essentially unknown. Therefore, in this study we aim to compare the changes in cerebral desaturation between different respiratory events in adult patients with SDB using NIRS. We hypothesize that obstructive and central sleep apneas can lead to larger and more profound cerebral desaturations compared to sleep hypopneas, considering that the airflow only partially drops in hypopneas but totally stops in apneas. Since OSA events can lead to stronger negative ITP that can reduce cardiac stroke volume compared to CSA events (i.e., reduced systemic blood supply from the heart) ^11, 35^, we hypothesize that the changes in cerebral StO2 and BV may be stronger in OSA than in CSA events.

## 2. Materials and Methods

### 2.1 Patients

We screened 33 newly diagnosed patients from our sleep center with severe SDB as indexed by an apnea-hypopnea index (AHI) ≥ 30/h but without comorbidities of severe unstable coronary/cerebral artery diseases, severe arterial hypertension/hypotension, or severe respiratory diseases. In the first night all patients underwent video-PSG recording (Embla RemLogic, Natus Medical Incorporated, Tonawanda, NY, USA) for diagnosis. On next day when diagnoses of SDB was confirmed, continuous positive airway pressure (CPAP) therapy was recommended as a suitable treatment solution by clinicians. In the following night, the patients underwent stepwise CPAP (AirSense™10, ResMed) titration together with video-polysomnography (PSG) and cerebral NIRS recordings: 1-h baseline sleep without CPAP followed by stepwise increment of 1-cmH2O pressure per-hour starting from 5 to 8 cmH2O depending on the individuals. Then only patients had both central and obstructive sleep apneas during the first 1-h baseline sleep without CPAP were included in our analyses, which were 13 patients. The demographics of these 13 patients were shown in Table 1. All patients gave their written informed consent. This study was approved by the local ethical commission of Northwest Switzerland, and was in compliance with the declaration of Helsinki.

**Table 1.**
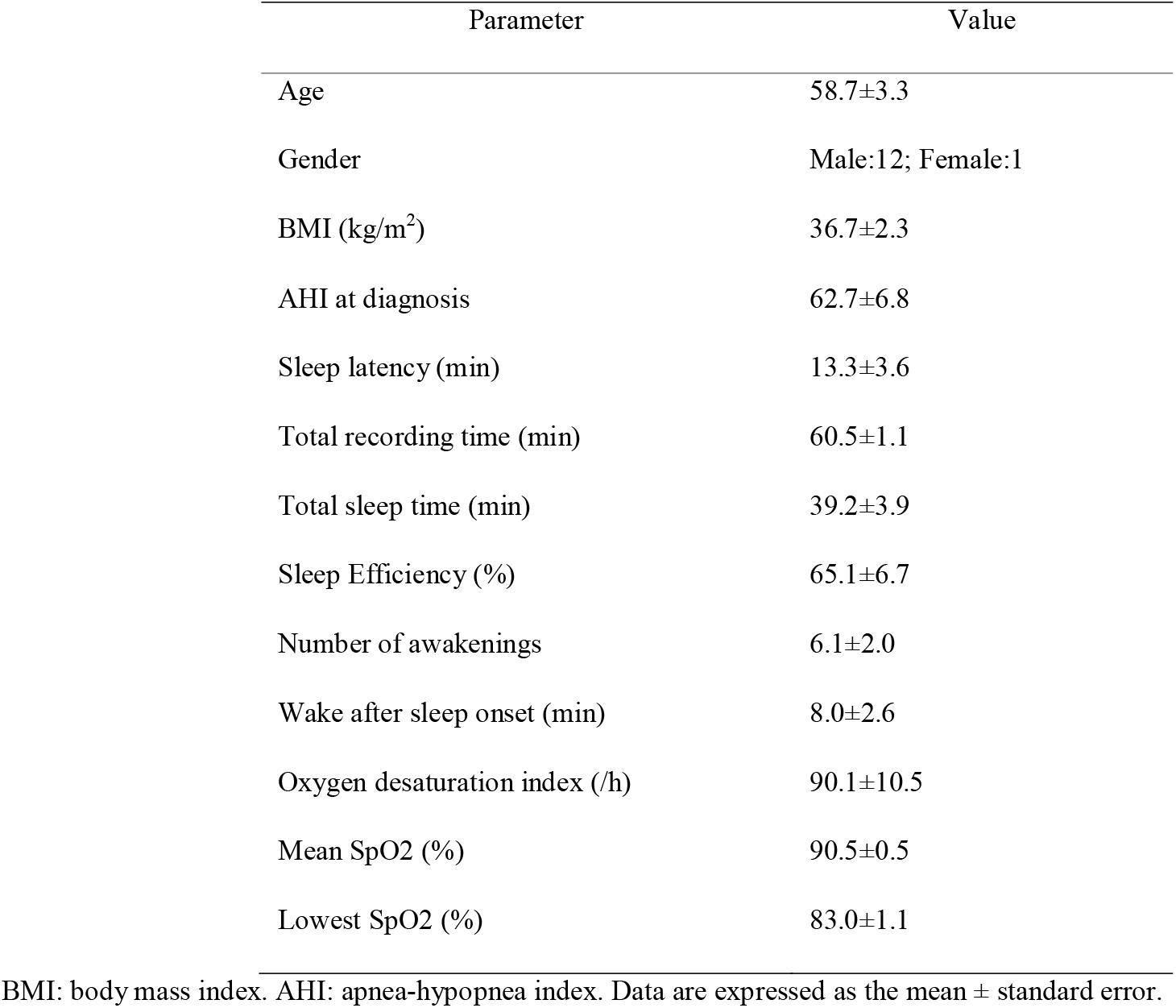
The demographics of patients and results of the first one-hour sleep polysomnography

### 2.2 Polysomnography

PSG comprehensively records the biophysiological signals during sleep. It includes electroencephalography (EEG) at electrode locations of F3, F4, C3, C4, O1, and O2 according to 10– 20 system, eye movements (electrooculogram, EOG), muscle activity (electromyogram, EMG), electrocardiogram (ECG), breathing functions (respiratory airflow and respiratory effort indicators), heart rate (HR) and peripheral SpO2. Each patient was videotaped with an infrared camera to allow for assessment of the movement during sleep. Two experienced sleep technologists independently scored the sleep stages, respiratory and limb movement events, and motion artifacts in 30-s epochs according to the 2017 American Academy of Sleep Medicine manual (AASM 2017^36^). The discrepancy between these two technologists was solved by their discussion or the recommendation of an experienced neurophysiologist. Only the data of the first 1-h recordings were used in this study.

### 2.3 Frequency-domain multi-distance near-infrared spectroscopy

Frequency-domain multi-distance (FDMD)-NIRS (Imagent, ISS, Champaign IL, USA) measurements were conducted over the middle of left forehead (i.e., in the middle of the area that below the hairline and above the left brow). The light emitters (8 laser diodes, 4 at 690 nm wavelength and 4 at 830 nm wavelength, so they were coupled into 4 sources) of the Imagent device were modulated at 110 MHz and the light can penetrate into the measured tissues with a depth of approximate 3-4 cm when the four coupled light sources were aligned and placed at 2 cm, 2.5 cm, 3 cm and 3.5 cm away from an optical fiber bundle that was connecting to the photomultiplier tube detector. The cerebral HbO_2_ and HHb were calculated from the fitted slopes of the modulated light amplitude and phase shifts over the four various distances^37, 38^. The sum of HbO_2_ and HHb (i.e., the total hemoglobin) was an index for the changes in cerebral BV. The absolute value of StO2 was also calculated in FDMD-NIRS^38^. Before the start of every patient recording the NIRS device was calibrated on optical phantom blocks.

The sample rate of FDMD-NIRS recording was 5.2 Hz. 2-s preceding and 5-s following the sleep apnea/hypopnea event was selected as baseline and recovery phase, respectively. The reliability of FDMD-NIRS measurement depended on the linearity of the raw optical signals on distances, i.e., the linear dependence *R*^*2*^ of modulated light amplitude and phase shift over the measure distances should be highly close to 1 in each light wavelength. The raw optical data were discarded if the *R*^*2*^ was smaller than 0.95 in either modulation amplitude or phase shift in any wavelength to exclude poor-quality data due to the improper probe-skin contact and shunted light reaching the detector without travelling through the tissue^17, 35, 39-41^. The StO2 values smaller than 30% or larger than 90% were discarded to exclude the potential movement artifacts or unreliable recordings, considering that the normal NIRS StO2 baseline value is between 50% and 80%^42, 43^. The NIRS data were subjected to a low-pass (<0.08 Hz) zero-phase filter designed using Hanning window to remove the physiological noises including heart rate, respiratory noise and spontaneous slow hemodynamic oscillations^17, 22, 35, 40, 41, 44^. Then the filtered data were smoothed with moving average smooth method (robust locally weighted scatter plot smoothing^45^ which assigns zero weight to data outside six mean absolute deviations)^17, 22, 35, 40, 41^. The coefficient of variation (CV, i.e., std./mean) of BV (CV-BV) during apnea/hypopnea and recovery phase was calculated. CV is a standardized measure of the variability of the data in relation to their mean, which can eliminate the bias due to individual differences. It has been used in previous NIRS studies to quantify the cerebral hemodynamic changes^28, 46^, including our previous study of cerebral BV changes in patients with OSA^17^. Smaller changes in CV-BV indicated smaller changes in the cerebral perfusion (i.e., the cerebral BV during the apnea/hypopnea event was more stable)^17^. The mean StO2 at baseline before event onset and the subsequent decrease (i.e., cerebral desaturation, which was the maximal value minus the minimal value after the event onset) were also calculated.

### 2.4 Statistical analyses

The changes in cerebral oxygenation can be influenced by many confounders (e.g., duration of event, severity of diseases, age, and sleep stages) besides the type of events. Therefore, linear mixed-effects model (LMM) with a random intercept by patients was used to quantify the differences in StO2 desaturation and CV-BV between hypopneas, OSA and CSA events respectively. Age, BMI, AHI, duration of respiratory event, mean HR during the event, and the baseline StO2 before the onset of the event were covariates added in the LMMs. Stepwise regression using backward elimination was done to automatically select the best explanatory variables by eliminating the non-significant covariates. Since there are different methods for calculating *R*^*2*^ for LMM to measure its goodness of fit, we reported both the conditional *R*^*2*^ calculated according to reference^47^ and Ω^*2*^ calculated according to reference^48^, which are two most frequently used *R*^*2*^, to assess the fitting quality of our final selected models^17, 41^.

Data were expressed as the mean ± standard error (SE) unless otherwise indicated. The pre-processing of NIRS signals were carried out in MATLAB (The Math-Works, Inc., Natick, MA, USA). All statistical analyses were performed using R (version 3.2.4). The LMM models were done using the R package *lme4*, and the automatic stepwise regression was done using the R package *lmerTest*.

## 3. Results

In total we successfully measured changes in NIRS signals during 277, 161 and 113 hypopneas, obstructive and central apneas during the one-hour baseline recordings, respectively. The sleep related parameters during this one-hour sleep are shown in Table 1. All respiratory events occurred during non-rapid eye movement (NREM) sleep, as none of the patients entered into REM sleep during their first hour sleep. The median of cerebral StO2 desaturation in CSA, OSA and hypopnea is 3.2% (interquartile range [IQR]: 2.3-4.8%), 2.4% (IQR: 1.9-3.0%) and 2.4% (IQR: 1.9-3.3%), respectively; and the corresponding median CV-BV is 1.3 (IQR: 1.1-1.7), 1.2 (IQR: 0.9-1.6) and 1.0 (IQR: 0.8-1.4), respectively.

It is not feasible to provide a figure showing the average changing trends of the NIRS signal during apnea/hypopnea events because of the large variations of the lengths of apnea/hypopnea events, i.e., the median duration of CSA, OSA and hypopnea is 17 seconds (IQR: 13-24 seconds), 18 seconds (IQR: 14-22 seconds) and 18 seconds (15-23 seconds), respectively. Thus a representative sample of changes in NIRS signals during different respiratory events is given in Fig.1. Cerebral HbO2 and StO2 decrease while HHb increases at event onset and all parameters show a reverse pattern after the recovery of breathing, which is a typical hemodynamic pattern triggered by sleep apneas and hypopneas as reported in previous studies^13,17-24^. Changes in cerebral BV are more heterogeneous. In some events BV first decreases and then increases during the events while a reversing pattern is seen in other events. These findings also fits the results reported in previous studies^22^. Although heterogenous these fluctuation of BV can still be interpreted as cerebral vasomotor activities^17^.

The results of the final LMM model selected by stepwise regression predicting changes in cerebral StO2 and CV-BV are summarized in Table 2 and Table 3, respectively. The conditional *R*^*2*^ and Ω^*2*^ of the model for StO2 are 0.77 and 0.67, respectively. These two values are 0.59 and 0.46 of the model for CV-BV, respectively. OSA and CSA events result in greater cerebral desaturations and larger fluctuations in cerebral BV compared to sleep hypopneas, but there is no significant difference between OSA and CSA events. The duration of the respiratory event is a significant predictor associated with changes in both the cerebral desaturation and BV. Events with higher baseline StO2 before their onsets have smaller desaturations during the events compared to the ones with lower baseline StO2. Higher mean HR during the event is associated with smaller changes in cerebral BV.

**Table 2.** The outcomes of the linear mixed-effects model of the cerebral StO2 desaturations.

| Covariates | Estimate<br>(10<sup>-1</sup>) | 95%CI<br>(10<sup>-1</sup>) | t-value | P-value |
| --- | --- | --- | --- | --- |
| Duration of event | 0.54 | [0.40, 0.68] | 7.59 | <0.0001 |
| StO<sub>2</sub> before event onset | -0.88 | [-1.35, -0.41] | -3.76 | 0.00021 |
| Types of events |  |  |  |  |
| OSA-CSA | -1.04 | [-3.69, 1.61] | -0.77 | 0.44 |
| OSA-Hypopnea | 1.78 | [-0.34, 3.90] | 1.65 | 0.098 |
| CSA-Hypopnea | 2.82 | [0.29, 5.35] | 2.18 | 0.029 |

CI: confidence interval. OSA: obstructive sleep apnea. CSA: central sleep apnea.

**Table 3.** The outcomes of the linear mixed-effects model of the changes in cerebral blood volume

| Covariates | Estimate<br>(10<sup>-1</sup>) | 95%CI<br>(10<sup>-1</sup>) | t-value | P-value |
| --- | --- | --- | --- | --- |
| Duration of event | 0.16 | [0.087, 0.23] | 4.17 | <0.0001 |
| Mean HR during event | -0.15 | [-0.22,-0.079] | -4.09 | <0.0001 |
| Types of events |  |  |  |  |
| OSA-CSA | 0.55 | [-0.84, 1.94] | 0.77 | 0.44 |
| OSA-Hypopnea | 2.96 | [1.84, 4.08] | 5.18 | <0.0001 |
| CSA-Hypopnea | 2.41 | [1.06, 3.76] | 3.50 | 0.0005 |

CI: confidence interval. HR: heart rate. OSA: obstructive sleep apnea. CSA: central sleep apnea.

## 4. Discussion

Our study for the first time compares the cerebral oxygenation between OSA and CSA events in adult patients with SDB. We find that CSA and OSA events cause similar cerebral desaturation and cerebral BV fluctuations, and these changes are greater in apneas compared to hypopneas. The similar hemodynamic response induced by OSA and CSA is remarkable considering the different pathomechanisms underlying both apnea events. OSA but not CSA causes a strong negative ITP which reduces the cardiac stroke volume and leads to an insufficient systemic blood supply. Such a decline in systematic hemodynamics imposes a stronger challenge for cerebral autoregulation in OSA compared to CSA. It is therefore reasonable to expect larger cerebral desaturations and BV fluctuations in OSA compared to CSA. However, our results contradict this hypothesis, suggesting that the cerebral autoregulation may be still capable to cope with the insufficient systemic blood supply due to negative ITP. Therefore, unlike the results previously published in children SDB^34^, CSA and OSA may have similar impact on cerebral oxygenation in adult patients even with severe SDB at least during the first hour of NREM sleep.

Another evidence supporting our assumption of functional cerebral autoregulation in our patients is the changing pattern of cerebral BV in OSA and CSA events shown in Fig. 1. Cerebral BV first decreases and later increases during later phase of the respiratory strains in both types of apnea events. This pattern has been previously reported in OSA events^13, 18, 22^. The increase in BV could be due to cerebral vasodilation which may be regulated by the cerebral autoregulation mechanisms. Further research is needed to test whether the increased cerebral BV during apneas is specific to intact cerebral autoregulation. We therefore suggest that future studies should focus on SBD-patients with impaired cerebral autoregulation such as patients with post stroke attack.

**Figure 1.**
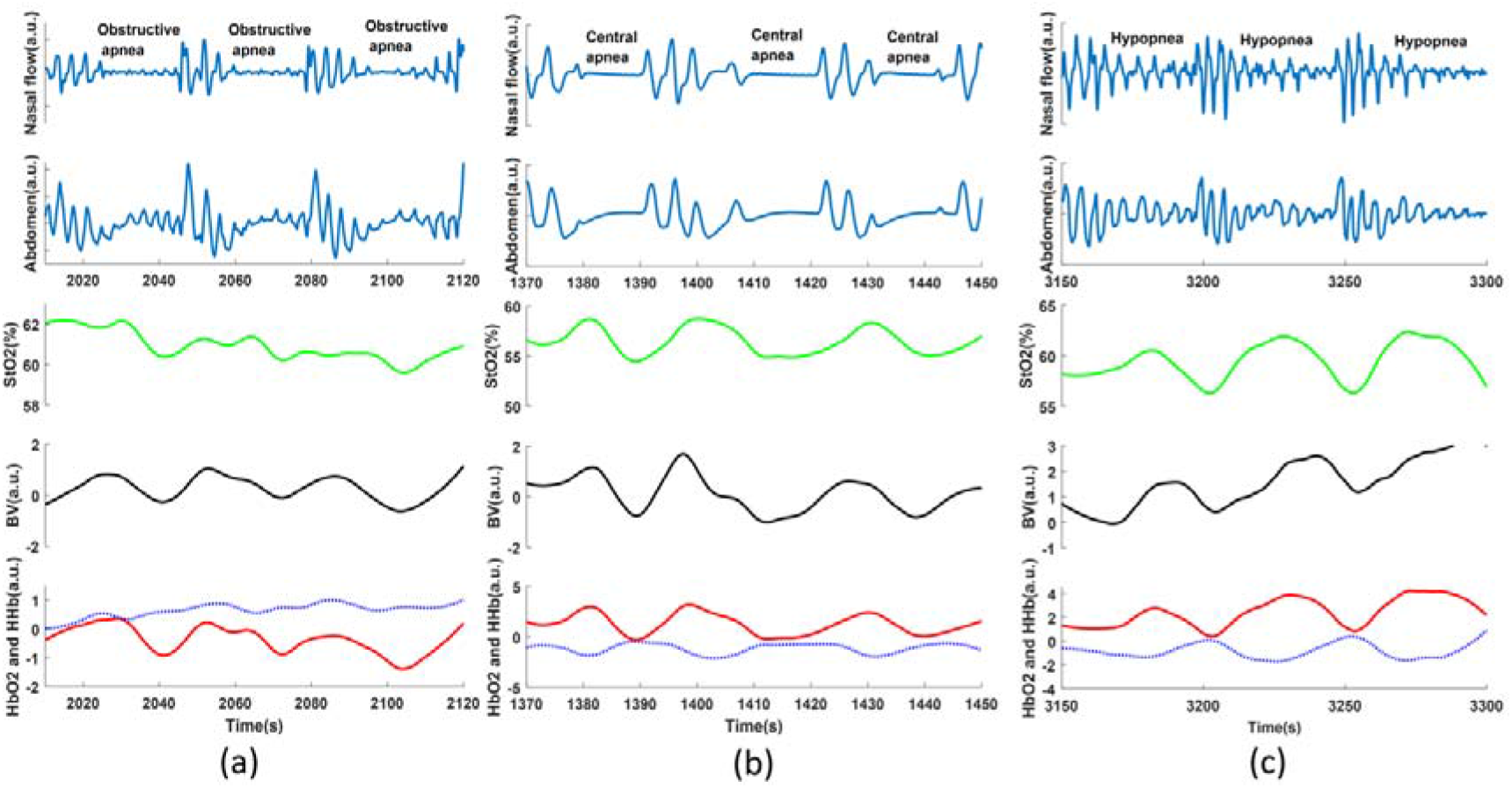
The samples of near-infrared spectroscopy (NIRS) changes in different respiratory events. BV is blood volume. a.u. is arbitrary unit.

In our results, longer respiratory events are associated with larger cerebral desaturation and changes in cerebral BV, which agree with the results of recent study comparing the cerebral oxygenations between obstructive sleep apneas and hypopneas^17^. These results are plausible because long cessation of inhaling causes longer period of oxygen deficit (i.e., greater hypoxia) thus resulting in greater cerebral desaturation. Longer pauses in exhaling can lead to larger accumulation of CO2 (i.e., greater hypercapnia) thus causing stronger cerebral vascular responses. Since longer apnea events are associated with larger decrease in HR^49^, it is reasonable that event duration and the mean HR during the event have contrary influences to the cerebral BV fluctuation (Table 3). Thus, our study directly provides evidences that longer respiratory events can cause more profound changes in cerebral hemodynamics. Currently, the clinical criteria of SDB severity solely rely on AHI. New metrics beyond the AHI criteria are desperately needed in this field^50^. We suggest that future studies should further investigate the relationship between longer respiratory events and the clinical outcomes (e.g., cardiovascular events including stroke) in patients with SDB, which may provide new metrics to better classify SDB.

There are two major limitations in our study. First, the data analyzed are only measured in the first hour of sleep without CPAP pressure, so all respiratory events are only from NREM sleep. We do not have data during REM sleep which mainly occurs in the later phase of sleep in the morning. This is because our data are part of another project aiming to characterize the cerebral hemodynamics during clinical routine CPAP titration. CPAP pressures during the titration can influence cerebral oxygenation^17^, or even trigger the so called treatment-emergent CSA in which the pathophysiological mechanism is still unclear^51^. Previous studies have shown that obstructive apneas/hypopneas are longer and associated with more severe SpO2 desaturations in REM sleep compared to NREM sleep^52-54^. Thus, although our selection of data only from the first hour of sleep ‘accidentally’ controlled for the confounder sleep stages, future studies comparing the impact of OSA and CSA on cerebral oxygenation during REM sleep are needed. Second, we have a relatively small sample size of 13 patients. Future studies with a large sample size including patients with different severities may provide better insights into the degree of cerebral oxygenation triggered by different respiratory events.

## Data Availability

All data produced in the present study are available upon reasonable request to the authors

## Acknowledgments

This work was supported by Clinic Barmelweid Scientific Foundation. The data acquisition work was supported by the Research Fund of the Swiss Lung Association No. 2014-22.

